# Barriers and Facilitators to the Implementation of Artificial Intelligence Enabled Diabetes Interventions in Lower-Middle-Income Countries: A Systematic Review Protocol

**DOI:** 10.1101/2025.06.01.25328752

**Authors:** Heya Desai, Tarun Katapally

## Abstract

**Background:** Diabetes represents an emerging global health crisis, with lower-middle-income countries experiencing a fast growth in prevalence. Diabetes care in these regions often faces significant challenges, including inadequate healthcare infrastructure, limited financial resources, a shortage of trained healthcare personnel, and a dual burden of communicable and non-communicable diseases. Artificial intelligence (AI) offers tailored and scalable solutions for addressing these systemic barriers by enabling early diagnosis and risk prediction, integrating diabetes care delivery across all levels of healthcare. However, the successful implementation of AI interventions requires an understanding of the unique infrastructural, technological, socio-political, and cultural factors influencing diabetes care in these regions.

**Objective:** This protocol outlines a systematic review to synthesize evidence on the barriers and facilitators to implementing AI-enabled diabetes interventions in lower-middle-income countries, and to examine the specific AI applications being deployed in these settings.

**Methods:** A comprehensive literature search will be performed across five databases: Medline, Web of Science, CINAHL, IEEE Xplore, and ACM Digital Library, encompassing peer-reviewed publications from 2015 to 2025. The review will include studies which assess AI-enabled interventions implemented in healthcare settings for diabetes prevention, diagnosis, and management in lower-middle-income countries. Studies that assess the efficacy of artificial intelligence tools without direct evaluation of these tools in clinical decision-making or patient care processes will be excluded. Two independent reviewers will assess studies for inclusion using a predefined search strategy. The reporting of results will adhere to the PRISMA 2020 checklist (1). The risk of bias in individual studies will be evaluated using Hawker’s tool for disparate study designs.

**Conclusion:** The findings of this systematic review will identify key considerations for implementing AI technologies in diabetes care and provide evidence to support policymakers, healthcare providers, and technology developers in designing context-appropriate interventions that improve care delivery and health outcomes in lower-middle-income countries.

## Introduction

Diabetes mellitus, encompassing type 1 (T1DM), type 2 (T2DM), and gestational diabetes (GDM), has emerged as one of the fastest-growing global health emergencies of the 21st century, with the prevalence of the non-communicable disease (NCD) rising from 200 million cases in 1990 to 830 million worldwide in 2022 (2). Diabetes and its related complications also significantly elevate the risk of developing other NCDs, including cardiovascular disease (3,4), stroke (5,6), and kidney disease (7,8). Since 2000, diabetes-related mortality has been on the rise globally (9), with an estimated 3.4 million deaths attributed to the condition and its complications in 2024 (10).

LMICs experience a disproportionate share of the global burden of diabetes mellitus, with approximately 80% of individuals with T2DM living in these regions (11). The rise in diabetes prevalence in LMICs, that of specifically T2DM and GDM continues to outpace that of high-income countries (HICs) (12) - a trend associated with rapid urbanization, sedentary lifestyles, nutritional transitions, and increasing obesity rates (13,14). While the incidence of T1DM is higher in HICs compared to LMICs (15), significantly higher cause-specific mortality rates are observed in LMICs (16). This pattern may be attributable to disparities in access to medications, inpatient and outpatient care, gaps in clinical guidelines, and treatment continuity (17–19) - with fewer than 10% of individuals with diabetes mellitus in LMICs receiving all guideline-recommended treatments for diabetes (20).

A subset of LMICs are lower-middle-income countries, defined by a Gross National Income per capita between $1,146 and $4,515 by the World Bank (21). In these settings, the prevalence of T2DM was the highest among all World Bank income classifications in 2021 (22), and over one-third of the population is at high risk of developing T2DM (23). However, prevention efforts—such as blood glucose screening, and physical activity and dietary counseling focused on fruits and vegetables—are limited (23)— and treatment coverage remains low (20,24).

Diabetes prevention and management in lower-middle income countries presents several challenges across sociodemographic, economic, infrastructural, and healthcare system domains (24–26). At the system level, the rise in diabetes occurs alongside an ongoing epidemiologic transition, where non-communicable diseases are increasing while infectious diseases remain prevalent (27,28). This results in health systems that are often designed to address acute conditions rather than provide long-term care for chronic diseases including diabetes (29,30). Clinical practice guidelines for diabetes mellitus (DM) management implemented in these regions rank lower in quantity and quality than those in high-income countries (HICs) and existing gaps in their content and implementation challenges may contribute to the higher diabetes mortality observed in these regions. For example, DM guidelines in lower-middle-income countries are often provider-focused, with limited emphasis on patient- and population-centered care, and do not cover the full spectrum of diabetes management. (31). Limited trained personnel and specialized care, along with centralized services in urban hospitals, leave rural and primary care facilities under-equipped for long-term management (30,32,33). Globally, lower educational attainment is associated with higher T2DM risk (34,35), and in lower-middle-income countries, inadequate access to diabetes education and counseling may heighten this risk by contributing to poor health literacy and limiting independent disease management (36–38).

Financial barriers, including a lack of health insurance (39,40), high out-of-pocket costs for medications, diagnostic tests, transportation to diabetes services, and consultations, further limit access to timely and continuous treatment (41,42). These challenges collectively contribute to delayed diagnosis, poor disease management, and preventable complications (43,44), highlighting the need for tailored strategies that consider local health system capacity and resource availability to improve access to care and diabetes outcomes in lower-middle-income countries.

Artificial intelligence (AI) is being increasingly integrated into tools and devices across the diabetes care continuum, offering benefits to healthcare providers, patients, and healthcare systems (45,46). AI systems can integrate diverse datasets—including electronic health records, continuous glucose monitoring, retinal images, genomics, wearable sensors, and behavioral data (e.g., physical activity and dietary habits)—to detect complex patterns, predict future risks (such as hypoglycemia and hyperglycemia), and support personalized treatment plans (47,48). In lower-middle-income countries, AI-enabled diabetes interventions can address access barriers and resource constraints, scaling healthcare capacity by assisting with tasks that may limit the reach of medical workforces. In the case of diabetic retinopathy (DR) care, AI algorithms can help improve efficiency, increase accessibility and reduce costs associated with existing tele-ophthalmology screening programs by autonomously screening for the condition (49,50). This tool can enable healthcare providers to extend DR assessments to remote areas, facilitating earlier detection and reducing condition progression. Furthermore, AI-powered remote monitoring tools—such as continuous glucose monitors, and mobile health applications can enable healthcare providers to monitor patient conditions from any location, reducing the need for in-person visits (45). In regions with a shortage of trained healthcare providers, AI can assist clinicians—without formal diabetes training—by analyzing patient data and providing evidence-based recommendations for treatment or lifestyle modifications (51,52). In patient-facing applications, AI technologies empower patients to make informed decisions about disease management by providing real-time, personalized feedback on behaviors such as diet, activity levels, and insulin dosing adjustments (49,53–55). These self-management tools can provide individuals the flexibility to tailor their management approaches according to changes in daily routines, enhancing long-term disease control and reducing the risk of complications.

Integrating AI into diabetes care in lower-middle-income countries has the potential to shift health systems toward more affordable, accessible, and personalized models of care, extending care to more individuals and enhancing treatment effectiveness and continuity (56–58).

As most available evidence on AI technologies for diabetes care originates from studies conducted in high-income settings (49,59), previous reviews have largely focused on these contexts. However, digital infrastructure (60,61), health system organization (62,63), clinical treatment targets (64,65), and the epidemiology of diabetes conditions (66,67) – including variations in risk factors (68) - often differ between high-income and lower-middle-income countries. These differences may influence regional implementation considerations that are important to address when applying AI-enabled interventions in lower-middle-income settings (58,69,70). While some reviews have explored the use of AI in other health domains in LMICs, such as dentistry (71), and others have assessed AI tools for diabetic retinopathy screening (57,58), no review to date has examined the barriers and facilitators to implementing AI interventions across diabetes care in lower-middle income countries. Addressing this gap may support the development and scale-up of contextually appropriate AI-enabled interventions across different levels of local health systems (69,72). Therefore, this systematic review aims to identify and synthesize evidence on the barriers and facilitators influencing the implementation of AI-enabled diabetes interventions in LMICs. Given the diversity of AI applications in diabetes care and the influence of local context (49,73), this review also aims to explore the specific AI applications and technologies being deployed in these settings and assess how their use aligns with local healthcare priorities, available infrastructure, and care delivery models.

## Methods

### 2.1 Design

This systematic review protocol follows the Preferred Reporting Items for Systematic Review and Meta-Analysis Protocols (PRISMA-P) 2015 guidelines (S1 Fig) (74). The primary research question guiding this review is ‘What are the barriers and facilitators to the implementation of AI-enabled diabetes interventions in lower-middle-income-countries?’. This protocol has been registered on the Open Science Framework and is available at: **osf.io/sq7gk**.

### 2.2 Eligibility Criteria

The inclusion and exclusion criteria for this review are summarized in Table 1. Only peer-reviewed articles published in English between 2015 and 2025 will be eligible for inclusion. Eligible studies must examine the implementation of interventions that use artificial intelligence (AI) technologies to support the screening, prevention, diagnosis, or management of diabetes, and report on physical and/or perceptual barriers and/or facilitators to their use. Eligible populations include individuals residing in LMICs who have been diagnosed with pre-diabetes— based on criteria established by the American Diabetes Association (75)—as well as those with T1DM, T2DM, and GDM. Studies focusing on healthcare provider experiences are included if the AI intervention was implemented in the context of delivering care to these populations. In this review, AI technologies are defined based on the World Health Organization’s definition as systems and tools that integrate algorithms capable of learning from data, enabling them to perform automated tasks without the need for explicit human programming at every step (76). LMICs are classified according to the World Bank’s fiscal year 2025 definitions. For the purposes of this review, the term “country” follows the World Bank’s usage, which refers to any territory for which authorities report separate social or economic statistics and does not necessarily indicate political independence. (21) As digital health interventions such as mHealth and eHealth tools may incorporate AI technologies, studies involving these tools will be included at the title and abstract screening stage for full-text review. During full-text assessment, studies will be excluded if the interventions do not involve AI technologies in their development or function; the presence of AI components will be confirmed at this stage. To ensure that the review captures real-world implementation in LMIC populations, only studies conducted within clinical, or healthcare settings will be included. Studies solely examining AI algorithm development, validation, or retrospective evaluation using population-level datasets, without evidence of real-world implementation or clinical application, will be excluded.

**Table 1.** Inclusion and exclusion criteria.

| **Criteria** | **Inclusion** | **Exclusion** |
| --- | --- | --- |
| **Geographical Scope** | Studies conducted in lower-middle-income countries according to the World Bank classification for fiscal year 2025. | Studies not conducted in LMICs or without a clearly defined geographic context. |
| **Language** | English | Non-English |
| Study Design | Qualitative, quantitative, or mixed-methods studies examining the implementation of AI-enabled interventions in lower-middle-income countries. | Theoretical or conceptual papers with no empirical data; purely descriptive studies not assessing implementation. |
| Implementation Focus | Studies that evaluate the implementation, adoption, integration, or user interaction of AI-enabled interventions in real-world settings. | Studies focusing only on model development, validation, or performance testing without deployment in a real-world setting. |
| Population | Studies involving human participants, including individuals with pre-diabetes or diagnosed with diabetes, healthcare providers, health system stakeholders involved in the delivery or implementation of AI interventions. | Animal studies, simulation-only studies without involvement of human users, patients, or healthcare providers in testing or evaluating the intervention. |
| **Condition Focus** | Pre-diabetes, type 1, type 2, and gestational diabetes. | Studies focusing exclusively on non-diabetic populations or on conditions not linked to diabetes prevention or care. |
|  |  | Studies where diabetes is incidental and not the primary focus of the AI intervention. |
| **Interventions** | Interventions using artificial intelligence (AI) technologies, including digital health tools that explicitly mention or apply AI methods in the intervention. | Interventions that do not use or mention AI technologies. |
| **Publication Type** | Peer-reviewed original research articles reporting primary data. | Editorials, commentaries, letters, opinion pieces, review articles (systematic, narrative, meta-analysis), conference abstracts or proceedings, study protocols, grey literature. |
| **Publication Status** | Published |  |
| **Publication Date** | 2015-2025 | Outside of this date range |
| **Outcomes** | Reports on barriers and/or facilitators to implementing AI-enabled interventions in diabetes care. | Studies that do not report any implementation-related outcomes. |
|  | Describes implementation outcomes such as adoption, acceptability, feasibility, integration, or sustainability. |  |

### 2.3 Data Sources

The review will include the following databases: Medline, Web of Science, CINAHL, IEEE Explore, and Association for Computing Machinery Digital Library (ACM Digital Library). Medline will be used to identify clinical and biomedical studies related to AI applications in diabetes care (77), while Web of Science offers multidisciplinary scientific research (78). IEEE Xplore and the ACM Digital Library will capture scientific, technical, and computing-focused literature (79,80).CINAHL will be used to access studies from nursing and allied health disciplines (81).

### 2.4 Search Strategy

Table 2 presents the search strategy for this review which focuses on four key themes: diabetes (82,83), artificial intelligence (82,84–86), digital health technologies(87–89), and lower-middle-income countries [refs]. Relevant terms for each theme were derived from previous literature on diabetes, artificial intelligence applications and digital health technologies within the context of diabetes care, as well as from systematic reviews conducted in LMIC and lower-middle-income country settings. The most recent database search was conducted on May 28, 2025. To identify any additional relevant studies not captured by the initial search strategy, the reference lists of peer-reviewed articles will also be reviewed.

**Table 2.**
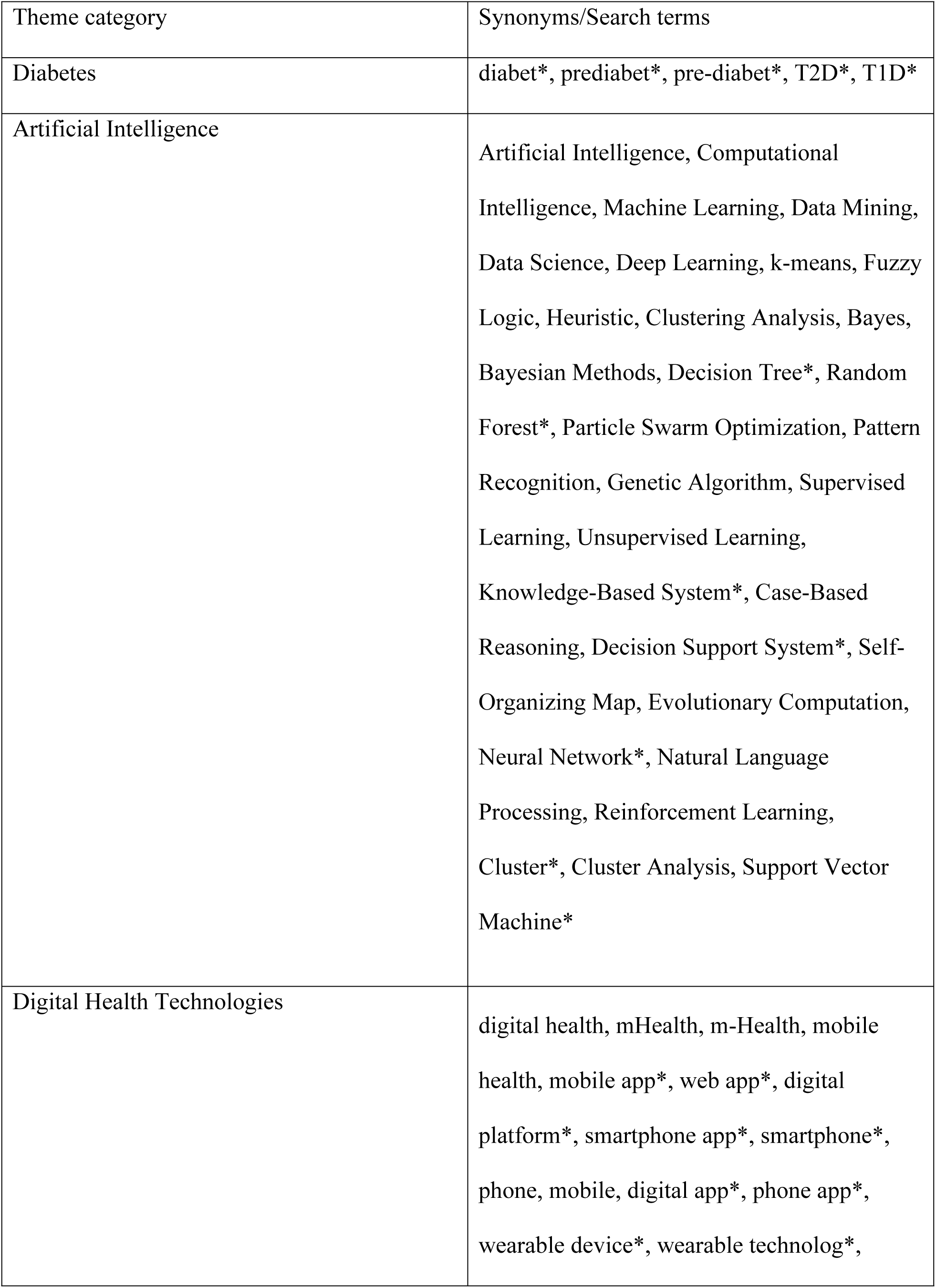
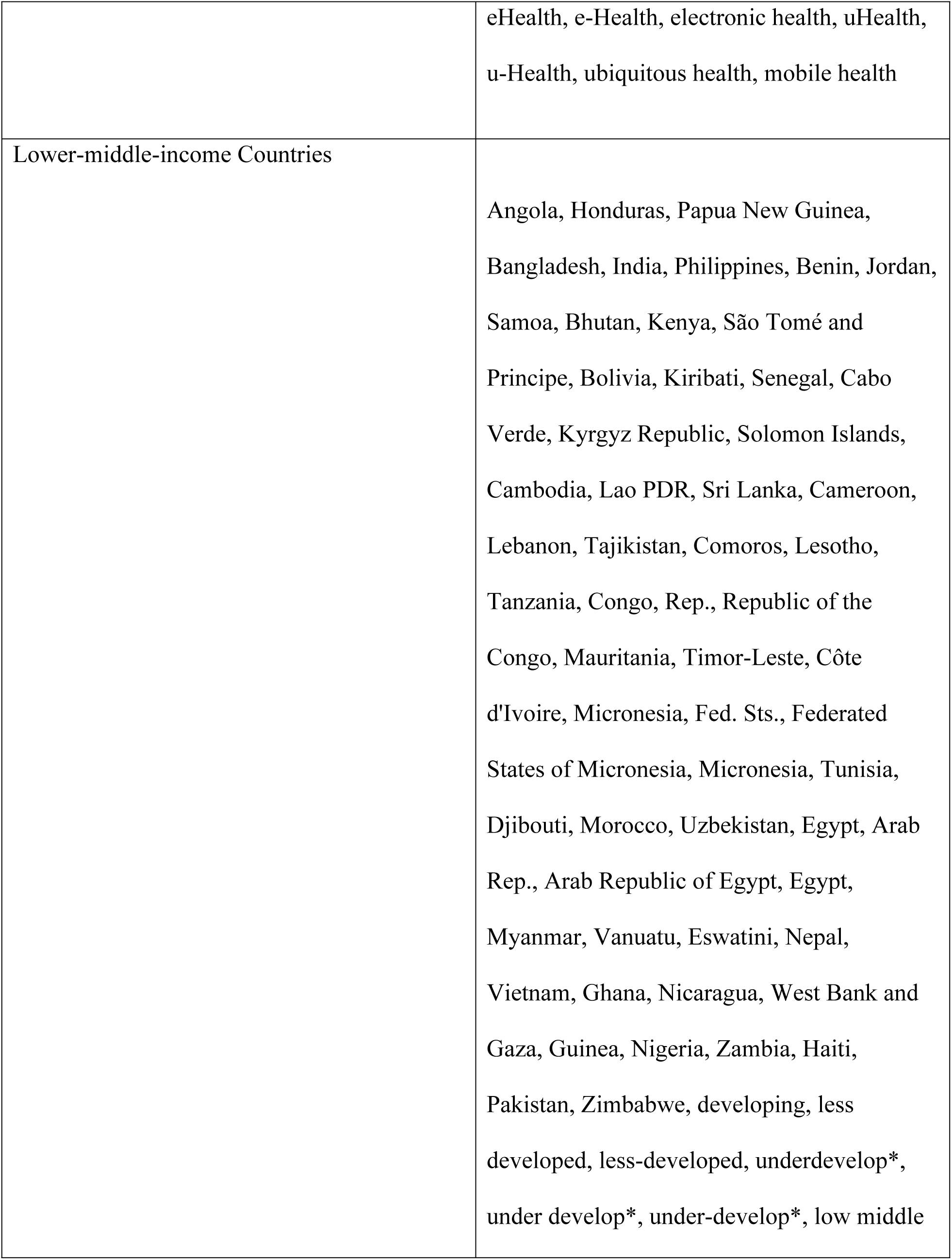
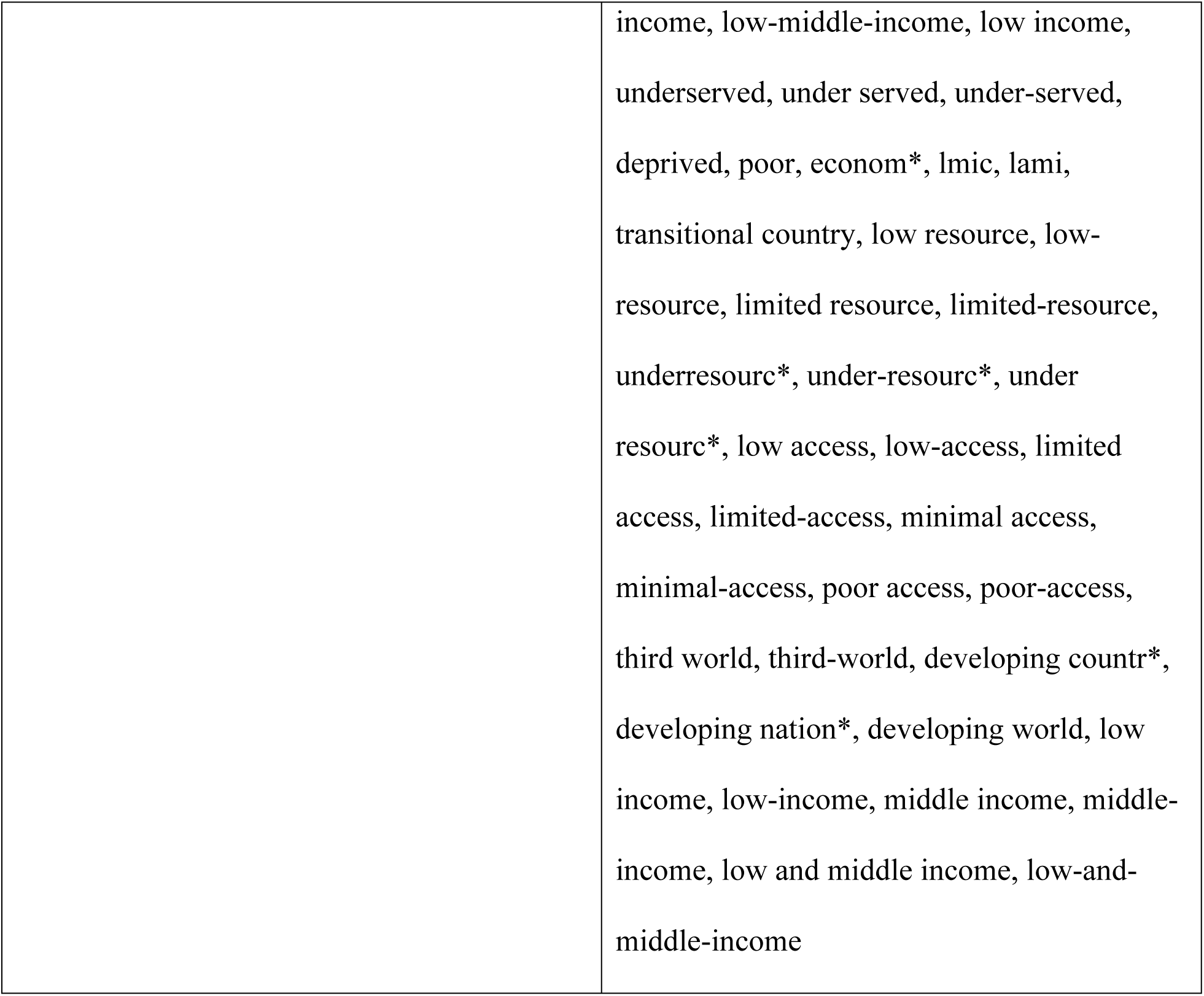
Key concepts and associated search terms used in the search strategy for this review.

### 2.5 Article Screening and Data Extraction

All search results from each database will be imported into the Covidence web-based platform for systematic reviews, to conduct screening and for reference management, including deduplication. Two independent reviewers will first screen titles and abstracts of all studies remaining after deduplication. Full-text screening will then be conducted, with any disagreements resolved through reviewer consensus. Data extraction will be performed independently by two reviewers using a standardized extraction form. Extracted information will include study characteristics (year of publication, study setting, study design, sample size, data sources, data collection period, eligibility criteria, study objectives, health system level (primary, secondary, tertiary)), population characteristics (population type (e.g., patient vs provider), sociodemographic details, diabetes type/risk status addressed, comorbidities), and intervention details (delivery model (e.g., clinic-based, mobile-based), intervention components, devices used, AI technology used, AI function and application context, implementation strategy).

Outcomes of interest will include health outcome measures, key results, and reported barriers and facilitators to implementation. Risk of bias will be assessed at the study level using the Hawker et al. (2002) checklist, which evaluates the methodological quality of qualitative, quantitative, and mixed-methods studies and is well suited to the scope of this review. The Mixed Methods Appraisal Tool (MMAT), which allows for the evaluation of methodological quality across diverse study designs, will be used to assess the strength of the body of evidence.

### 2.6 Limitations

This review is limited to studies conducted in lower-middle-income countries, which facilitates a targeted synthesis of evidence within a specific economic and health system context, however, may exclude relevant insights from middle and upper-middle-income countries that may experience similar barriers and facilitators implementation challenges. The variation between AI technologies, implementation strategies, and health systems infrastructures across lower-middle-income countries may introduce significant heterogeneity, limiting the comparability of findings. To account for contextual variability, key study setting variables (e.g., country, care level, delivery model) will be extracted and findings will be organized by geographic region and population type to identify patterns in implementation barriers and facilitators that may be influenced by local health system structures and care delivery settings. Furthermore, limiting the review to digital health interventions that explicitly incorporate AI excludes non-AI-based tools used across the diabetes care continuum in lower-middle-income countries. This focus may limit the generalizability of findings to implementation considerations for broader digital health interventions; however, it addresses a gap in the evidence by identifying distinct challenges and opportunities associated with data-driven AI-enabled tools.

## 3 Conclusion

This systematic review will examine the implementation of AI-enabled diabetes interventions in lower-middle-income countries. By examining reported barriers, facilitators, intervention types, and AI applications, this review will identify implementation considerations to support policymakers, healthcare providers, and technology developers in advancing the design and integration of AI-enabled tools for diabetes care. The review also aims to highlight evidence gaps and priority areas for future research to enhance the feasibility, scalability, and uptake of interventions that enable proactive, personalized, and locally responsive care aligned with healthcare system needs and available resources.

## Data Availability

No datasets were generated or analysed during the current study. All relevant data from this study will be made available upon study completion.

## Acknowledgements

This research was funded by the Canada Research Chair which supports Dr. Tarun Katapally’s research programme.

## Author Contributions

**Heya Desai:** contributed to conceptualization, investigation, and writing-original draft preparation, review and editing. **Tarun Reddy Katapally:** contributed to conceptualization, investigation, funding acquisition, supervision, and writing-review and editing.

---

**S1 Fig.** PRISMA-P (Preferred Reporting Items for Systematic review and Meta-Analysis Protocols) 2015 checklist: recommended items to address in a systematic review protocol.

